# Cost-effectiveness of amyloid-targeting therapies in a memory clinic population; a simulation study

**DOI:** 10.64898/2026.07.30.26359218

**Authors:** P.J. van der Veere, H.M. Broulikova, R. Handels, C.E. Teunissen, L.E. Collij, E.G.B. Vijverberg, W.M. van der Flier, J. Berkhof

## Abstract

**Objectives:** The cost-effectiveness of new amyloid-targeting therapies (ATTs) for patients with mild cognitive impairment (MCI) or mild dementia due to Alzheimer’s disease (AD) is influenced by assumptions about treatment effectiveness beyond the trial durations. To assess the cost-effectiveness of ATTs over a lifetime horizon, an AD microsimulation model was applied.

**Methods:** The AD microsimulation model is based on statistical joint models, which link cognitive decline (Mini-Mental State Examination [MMSE]) and states of functional independence (MCI, dementia, institutionalisation), fitted to the Amsterdam Dementia Cohort. The time from MCI to death was simulated under care-as-usual (CAU) and two ATT scenarios, assuming an ATT duration of eighteen months and treatment effect waning of 0% (no-waning) or 20% per year. The main outcome was the incremental cost-effectiveness ratio (ICER), defined as incremental costs per quality-adjusted life year (QALY) gained. A societal perspective was taken for costs and effects.

**Results:** The ATT scenario without waning resulted in 0.72 additional QALYs and €28,502 additional costs per person compared to CAU. At a list price of €22,600/year for the ATT, the ICER was €39,745/QALY for the no-waning and €151,016/QALY for the 20%-waning scenario. At a willingness-to-pay threshold of €20,000/QALY, the corresponding threshold prices were €10,500 (95% CI: <€0 to €23,000; no-waning) and <€0 (95% CI: <€0 to €2,250; 20%-waning), respectively.

**Conclusions:** The cost-effectiveness of ATTs is strongly influenced by the waning of the treatment effect. A favourable cost-effectiveness profile was only achieved when the treatment effect did not wane after the eighteen-month treatment period.

## Introduction

Alzheimer’s disease (AD) is the most common cause of dementia, a leading cause of reduced quality of life globally and a major driver of healthcare costs.^1, 2^ As the number of older adults rises,^3^ the burden of dementia on society through formal and informal care will likewise increase in the coming decades. Since the 2000s, cholinesterase inhibitors have been available for dementia patients with a clinical phenotype of AD, alleviating symptoms with an effect size equivalent to a few months of cognitive decline.^4^ Recently, amyloid-targeting therapies (ATTs) have gained market access, a new drug class with disease-modifying properties for patients with biomarker-confirmed AD. The first two drugs in this class, lecanemab and donanemab, slowed cognitive and functional decline in patients with mild cognitive impairment (MCI) and mild dementia due to AD by about 30% over an eighteen-month trial period.^5, 6^

While ATTs have gained market access in many countries, reimbursement of these therapies remains uncertain. This uncertainty stems from their modest clinical effect combined with their high list prices of $26,000 and $32,000 per year for, respectively, lecanemab and donanemab in the USA.^7, 8^ Furthermore, determining the value of the modest clinical effects is complicated by the short trial period of eighteen months, while clinical symptoms due to AD unfold over a decade.

The impact of treatment uncertainties on the costs and effects of ATTs can be explored using simulation models. Several cost-effectiveness analyses of ATTs have been published, underscoring the importance of treatment effect waning and treatment duration on the cost-effectiveness.^9–19^ However, these simulation models are hampered by three limitations. First, most existing models assumed that ATTs directly altered the transition rates between discrete disease stages (MCI, mild dementia, moderate dementia, etc.).^9–16^ They did not account for gradual cognitive decline as the primary determinant of functional decline in AD,^20^ which matters because cognitive benefit may gradually build-up after the start of treatment, and cognition and functional independence are related but distinct concepts. Second, some of the existing models included patients with clinical diagnoses of MCI or mild dementia due to AD in the population used to develop the models,^14–19^ while there is substantial misclassification of AD by clinicians when biomarker confirmation is absent.^21^ Third, the simulation models were constructed using multiple data sources and transition probabilities reported in the literature,^9–19^ reducing their internal validity.

We have previously developed statistical joint models that can be used for microsimulation of health trajectories of individuals potentially eligible for ATTs, i.e. amyloid-positive MCI and dementia memory clinic patients.^22^ The joint models were estimated using data of participants of the Amsterdam Dementia Cohort and incorporated a link between cognitive decline on a continuous scale (Mini-Mental State Examination [MMSE]) and stages of functional independence (MCI, dementia, institutionalisation). The aim of this study was to estimate the cost-effectiveness of ATTs in a Dutch memory clinic population of amyloid-positive MCI and mild dementia patients. We focused on how assumptions about treatment effect waning, treatment duration, and the modelling of cognition and functional independence affected these estimates. Given this objective, lecanemab and donanemab were not evaluated individually but modelled as a single representative ATT treatment.

## Methods

### Model description

The AD microsimulation model was developed previously and is described in detail in the supplemental methods.^22^ A model flow chart is presented in Figure 1. The model was built from four statistical models: two joint models (model 1 and 2) and two Weibull models (model 3 and 4). Joint models combine longitudinal markers and a time-to-event model.^23^ Model 1 describes the longitudinal pattern of MMSE in the MCI stage and the transition from MCI to dementia in the community. The person-specific expected MMSE value was included as a time-varying variable in the time-to-event portion of the joint model. Model 2 describes the longitudinal pattern in MMSE in the dementia stage and the transition from dementia in the community to the competing events of institutionalisation and death outside an institution in the first four years after the dementia diagnosis. Again, the person-specific expected MMSE value was included as a time-varying variable in the time-to-event portion of the joint model. Model 3 is a Weibull model for the transition from dementia in the community to the competing events of institutionalisation and death outside an institution if they occurred more than four years after the dementia diagnosis. Events occurring before and after four years after the dementia diagnosis had to be modelled separately because MMSE measurements were only available for the first four years. Model 4 is a Weibull model for the time from institution to death. The transition from MCI to death was informed by Dutch age- and sex-specific life tables.

**Figure 1:**
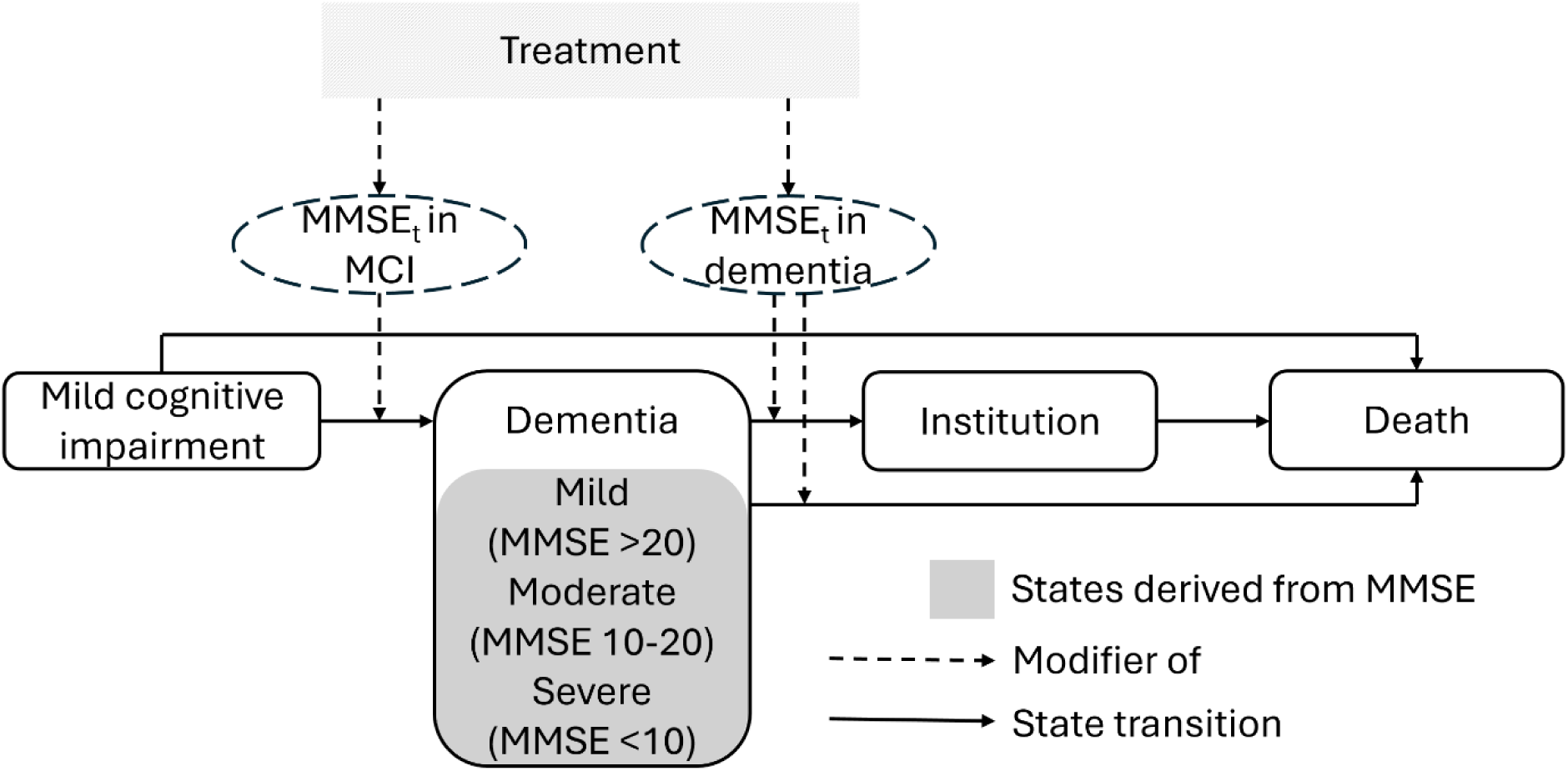
Graphical model overview. States are depicted by rectangular boxes. The oval MMSEt box depicts the longitudinal expected MMSE trajectory, modifying the transition from mild cognitive impairment to dementia, and from dementia to institution or death outside an institution. In the treatment scenarios, this MMSEt trajectory is modified by the treatment effect. Abbreviations: MMSE = Mini-Mental State Examination

People in the dementia in the community state can be in a mild, moderate, or severe dementia state. These states have different associated costs and utilities. The transition between the dementia in the community states is determined by the MMSE level with thresholds of 20 (from mild to moderate dementia) and 10 (from moderate to severe dementia).^24^ The time in each state is extracted from the models using inverse transform sampling.

### Simulation inputs

Input parameters and data sources are shown in Table 1.

**Table 1:** Simulation inputs. * As the parameter uncertainty was unknown, 20% of the point estimate was taken as the bounds for a uniform distribution. Abbreviations: ADC = Amsterdam Dementia Cohort, MCI = Mild Cognitive Impairment

| Parameters | Value | Uncertainty range | Distribution | Source |
| --- | --- | --- | --- | --- |
| **General** |  |  |  |  |
| Annual discount rate for costs | 3 | - | Fixed | Dutch Costing Manual<sup>33</sup> |
| Annual discount rate for utilities | 1.5 | - | Fixed | Dutch Costing Manual<sup>33</sup> |
| Proportion of simulation cohort starting with MCI | 89.1% |  | Fixed | Claus et al. (2025)<sup>25</sup> |
| **Baseline characteristics MCI due to AD cohort** |  |  |  |  |
| Age, mean ± SD years | 66 ± 7 | - | Bootstrapped | ADC<sup>26</sup> |
| Female | 44% | - | Bootstrapped | ADC<sup>26</sup> |
| MMSE at baseline, mean ± SD | 26 ± 2 | - | Bootstrapped | ADC<sup>26</sup> |
| **Baseline characteristics mild dementia due to AD cohort** |  |  |  |  |
| Age, mean ± SD years | 66 ± 7 | - | Bootstrapped | ADC<sup>26</sup> |
| Female | 51% | - | Bootstrapped | ADC<sup>26</sup> |
| MMSE at baseline, mean ± SD | 24 ± 3 | - | Bootstrapped | ADC<sup>26</sup> |
| **Treatment effects** |  |  |  |  |
| Slowing of speed of cognitive decline | 30% | 22% to 38% | Normal | See supplementary method for uncertainty estimation<sup>5, 6</sup> |
| ARIA probability, %/year | 20% | - | Fixed | See supplementary methods<sup>27</sup> |
| Proportion of ARIA that were symptomatic | 22% | - | Fixed | See supplementary methods<sup>27</sup> |
| Reduction in ARIA probability after 18 months, %/year | 100% | - | Fixed | Assumed |
| Aria duration, months | 3 | - | Fixed | Assumed |
| Proportion of treatment eligibility in diagnosed patients | 15% | - | Fixed | Claus et al. (2025)<sup>25</sup> |
| Time to clear amyloid plaques, after which treatment stop | 18 months | - | Fixed | Assumed |
| Assumptions regarding treatment effects |  |  |  |  |
| Voluntary discontinuation, %/year | 0 | - | Fixed | Assumed |
| Effect waning, %/year | 0%, 20% | - | Fixed | Assumed |
| **Cost** |  |  |  |  |
| Stages, per year |  |  |  |  |
| MCI | 12,379 | 7,718 to 17,039 | Normal | See supplementary methods<sup>33, 34</sup> |
| Mild Dementia | 30,977 | 22,636 to 39,318 | Normal | See supplementary methods.<sup>33-35</sup> |
| Moderate dementia | 46,006 | 36,036 to 55,976 | Normal | See supplementary methods.<sup>33-35</sup> |
| Severe Dementia | 77,186 | 53,930 to 100,441 | Normal | See supplementary methods.<sup>33-35</sup> |
| Institutionalisation | 110,688 | - | Fixed | See supplementary methods.<sup>33-35</sup> |
| Treatment related |  |  |  |  |
| Medication costs | 22,600 | - | Fixed | Lecanemab list price<sup>7, 32</sup> |
| Infusion cost | 171 | - | Fixed | Dutch Costing manual<sup>33</sup> |
| MRI | 374 | - | Fixed | See supplementary methods<sup>33</sup> |
| Treatment of symptomatic severe ARIA treatment, per episode | 3,970 | - | Fixed | See supplementary methods<sup>33, 43</sup> |
| Diagnostic costs memory clinic | 523 | - | Fixed | See supplementary methods<sup>33</sup> |
| Additional diagnostics to determine treatment eligibility | 542 | - | Fixed | See supplementary methods<sup>44, 45</sup> |
| **Utilities** |  |  |  |  |
| Symptomatic ARIA disutility | -0.07 | -0.10 to -0.03 | Normal | Luenogo-Fernandez et al. (2013)<sup>31</sup> |
| For patients by stage |  |  |  |  |
| MCI | 0.80 | 0.75 to 0.85 | Normal | Landeiro et al. (2020)<sup>30</sup> |
| Mild Dementia | 0.74 | 0.69 to 0.79 | Normal | Landeiro et al. (2020)<sup>30</sup> |
| Moderate dementia | 0.59 | 0.47 to 0.71 | Normal | Landeiro et al. (2020)<sup>30</sup> |
| Severe Dementia | 0.36 | 0.18 to 0.53 | Normal | Landeiro et al. (2020)<sup>30</sup> |
| Institutionalised | 0.36 | 0.18 to 0.53 | Normal | Assumed same as for severe dementia |
| Disutility partner by stage |  |  |  |  |
| MCI | -0.03 | -0.36 to -0.024 | Uniform* | Lin et al. (2023)<sup>11</sup> |
| Mild dementia | -0.05 | -0.06 to -0.04 | Uniform* | Lin et al. (2023)<sup>11</sup> |
| Moderate dementia | -0.085 | -0.102 to -0.068 | Uniform* | Lin et al. (2023)<sup>11</sup> |
| Severe dementia | -0.10 | -0.12 to -0.08 | Uniform* | Lin et al. (2023)<sup>11</sup> |
| Institutionalised | -0.10 | -0.12 to -0.08 | Uniform* | Assumed same as for severe dementia |

#### Study population and setting

The simulated cohort consisted of 89.1% amyloid-positive patients with MCI and 8.9% amyloid-positive patients with mild dementia in the community.^25^ Patients were sampled from the population used to develop models 1 and 2, which were participants of the Dutch memory clinic Amsterdam Dementia Cohort.^26^ The MCI patients had a mean (SD) age of 66 (7) years, a mean (SD) MMSE of 26 (2), and 44% were female. The mild dementia patients had a mean (SD) age of 66 (7) years, a mean (SD) MMSE of 24 (3), and 51% were female.

#### Treatment implementation and side-effects

The randomized trials showed that treatment with ATTs slowed clinical disease progression by about 30% over eighteen months, as measured by continuous scales of cognition and functional independence.^5, 6^ Based on the conceptual model of AD, where cognitive decline leads to a loss of functional independence,^20^ we operationalised the treatment effect as a 30% slowing of MMSE decline (see supplemental figure 1 for MMSE curves and transition probability curves under treatment). We assumed treatment stopped after eighteen months, but effects continued while patients were in the community. All participants were assumed to be treatment adherent, without voluntary discontinuation.

During ATT treatment, the yearly probability of developing amyloid-related imaging abnormalities (ARIA) was 20% based on the reported incidence of ARIA in the lecanemab and donanemab trial for APOE ε4 non-carriers and heterozygotes (see supplemental methods and supplemental tables 1 and 2 for details).^27^ They were classified based on accompanying symptoms (yes, no) and radiological severity (mild, moderate, and severe). All ARIAs were assumed to last three months, with monthly MRIs until a scan without ARIA was obtained. Based on the appropriate use recommendations, monitoring for ARIA was performed at 8, 12, 28, and 52 weeks after treatment initiation.^28, 29^ Treatment was discontinued at an observed severe ARIA (4.1%) and at the second observed ARIA (1.8%). Treatment was suspended for the duration of symptomatic mild ARIA (4.3%) and observed moderate ARIA (6.1%).

#### Utilities and disutilities

The patient utilities of the clinical AD stages (MCI, mild, moderate, and severe dementia), based on caregiver-proxy valuations, were obtained from the literature.^30^ Patients were assumed to have one care partner, for whom state-specific disutilities were included.^11^ For symptomatic ARIA, a disutility of a transient ischemic attack (-0.07) was applied for three months.^31^

#### Treatment costs

The price of ATTs was set at €22,600 based on the price of lecanemab in the US and Germany.^7, 32^ The treatment costs included the cost of assessing ATT-eligibility based on cerebrospinal fluid biomarkers and APOE-genotyping, assuming a 15% eligibility probability (supplemental table 3);^25^ four-weekly infusion costs;^33^ monitoring for ARIA based on MRI and clinical assessments; and treatment for symptomatic severe ARIA (supplemental table 4). See supplemental methods for details.

#### Care Costs

Care costs per state included formal and informal care costs (supplemental table 5). Formal care costs for the clinical AD stages were based on a cost-of-care study of amyloid-positive patients in the Swedish national dementia registry.^34^ In the institutionalisation state, costs for housing and nursing were based on a price set by the Zorginstituut Nederland (ZiN; Dutch National Health Care Institute),^33^ and further formal care costs were sourced from a Swedish cost-of-care study.^34^ Informal care costs were included by multiplying the hours of informal care by the opportunity costs set by ZiN.^33^ The hours of informal care were based on own data for the MCI state (see supplemental methods) and a meta-analysis of multiple European cohorts for the dementia states.^35^

#### Currency, inflation correction and discounting

All prices were inflation-adjusted to represent 2022 euros.^36^ Future costs were discounted by 3% per year and future utilities by 1.5%.^37^

### Cost-effectiveness analysis

We compared the disease trajectory under two ATT scenarios against care-as-usual (CAU). In the no-waning scenario, the treatment effect remained stable when treatment provision stopped after eighteen months. In the waning scenario, the treatment effect waned by 20% per year when treatment provision stopped after eighteen months. Care-as-usual in AD currently consists of best supportive care.^38^

Model outcomes were costs and quality-adjusted life years (QALYs), a combination of quality of life and years lived, from a societal perspective.^37^ Model outcomes were compared over a lifetime horizon using incremental cost-effectiveness ratios (ICER) per QALY gained. Both ATT scenarios were compared to CAU at the ATT list price. In addition to the analyses at the list price, we calculated threshold prices for the ATTs at willingness-to-pay (WTP) thresholds ranging from €0 to €200,000.

Uncertainty around the model outcomes was estimated for the main analyses by varying the model parameters and the costs and utilities over 1000 parameter sets drawn from their respective distributions. The probability of the ATT-to-CAU comparisons meeting a €20,000/QALY WTP and €50,000/QALY threshold were calculated at the ATT list price. The €20,000/QALY threshold was chosen based on the Dutch WTP for diseases with a utility loss equivalent to MCI or mild dementia.^39^ The €50,000/QALY threshold was chosen based on the Dutch proposed WTP threshold for preventative interventions.^40^ The probabilities of the ATT-to-CAU comparisons meeting a WTP of 80,000/QALY was also calculated.

Simulated cohorts included 100,000 patients per scenario to ensure stable results, except for the cohorts used to estimate uncertainty around the outcomes of the main analyses. Due to computational limitations, these included 10,000 individuals per parameter set for each of the two ATT scenarios and CAU. Reporting of this study was performed in accordance with the Consolidated Health Economic Evaluation Reporting Standards (CHEERS) guidelines.^41^

### Sensitivity analysis

We determined the cost-effectiveness of ATT when starting treatment in either MCI or mild dementia, performed one- and two-way parameter sensitivity analysis starting from the no-waning scenario, and performed structural sensitivity analyses. See the supplementary Excel table for an overview of all sensitivity analyses.

Parameter sensitivity scenarios starting from the no-waning scenario were obtained by applying the following changes: (1) Treatment provision shortened to 0.5 years, after which treatment effects persisted while patients were in the community. (2) Treatment provision was extended to continue treatment until moderate dementia was reached (MMSE below 20), at which point provision and effects both stopped. (3) Treatment effect size was changed to 15% and 45% slowing of cognitive decline. (4) The treatment effect was stopped when moderate dementia was reached (MMSE below 20). (5) Voluntary discontinuation of treatment was set to 10% per year during the eighteen months of treatment provision, without enduring treatment effects after treatment discontinuation. (6) The cost of assessing ATT-eligibility was not included in the treatment costs. (7) The cost of infusion was not included in the treatment costs (analogous to a subcutaneous provision). (8) The discounting factors for costs and utilities were both set to 0% and to 5%. (9) A healthcare perspective was adopted by omitting informal care costs and partner disutilities.

To explore structural uncertainty, we compared our primary approach of slowing cognitive decline with two alternative operationalisations of the ATT effect. First, we assumed that ATTs had a direct effect on functional independence. For this scenario, we estimated Weibull hazard models for all state transitions except MCI to death (i.e. MCI to dementia, dementia to institution or death outside institution, and institution to death). In the ATT scenarios, the hazard of transitioning from MCI to dementia and dementia to institution or death outside of institution was reduced by 30%. In the dementia state, treatment effects were only applied to patients with mild dementia. See supplemental methods for further details. Second, we assumed an ‘all-or-nothing’ treatment effect, where 30% of the simulated patients remained cognitively stable (‘all’) and 70% experienced no treatment effect (‘nothing’, i.e. natural decline). This is an extreme example of treatment effect heterogeneity. In the other analyses, the ATT effect can be considered ‘leaky’, as everyone experiences a 30% slowing in cognitive decline.

Simulations were performed in R version 4.4.3.^42^ Code used in the simulations is available at https://github.com/PieterVeere/ADC_CEA_sim.

## Results

The ATT no-waning and waning scenarios had higher (discounted) QALYs and costs than CAU (Figure 2). Under CAU, the mean QALYs per person were 5.3 and the mean costs per person were €353,410. In the ATT no-waning scenario, the mean QALYs per person were 6.0 and the mean costs per person were €381,912. Compared to CAU, the increase in QALYs was 0.72 and the increase in costs was €28,502, yielding an ICER of €39,745/QALY. At a WTP of €20,000/QALY, there was a 3% probability that ATT was cost-effective (Supplemental Figure 2), while at WTPs of €50,000/QALY and €80,000/QALY the probability was, respectively, 68% and 98%.

**Figure 2:**
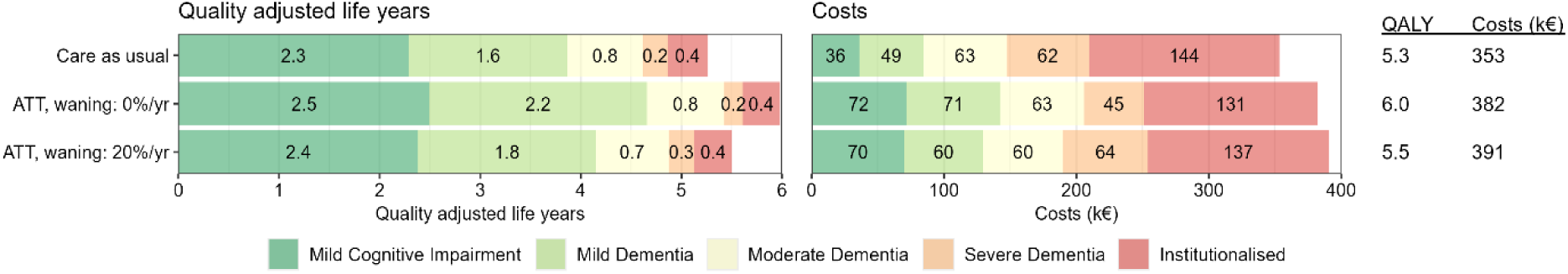
Quality adjusted life years and costs of the simulations by severity. Mean QALYs and costs per state are depicted within the bar. The cost of treatment were included in the state during which the treatment was provided. Abbreviations: QALY= quality adjusted life year; ICER = incremental cost-effectiveness ratio.

In the ATT waning scenario (20% treatment effect waning per year), the mean QALYs per person were 5.5 and the mean costs per person were €390,651 (Figure 2). Compared to CAU, the increase in QALYs was 0.25 QALYs and the increase in costs was €37,242, yielding an ICER of €151,016/QALY. At a WTP of €20,000 to €80,000/QALY, there was a 0% probability that ATT was cost-effective (Supplemental Figure 2). For both treatment scenarios, additional QALYs and costs were seen in the MCI and mild dementia stages (Figure 2).

In the ATT scenarios, ARIA-related costs were driven by MRI monitoring costs. The mean lifetime costs per person related to ARIA were €1,703. Due to the low frequency of symptomatic ARIA, the mean loss in QALY per person due to ARIA was -0.001.

The price at which ATT was cost-effective changed strongly if treatment effect waning was assumed (Figure 3). In the no-waning scenario, the threshold drug price was €10,500/year (95% confidence interval [CI]: <€0 to €23,000) at a WTP of €20,000/QALY, and €25,750/year (95% CI: €11,750 to €44,750) at a WTP of €50,000/QALY. This corresponds to a total treatment drug costs of, respectively, €15,750 and €38,625 for eighteen months of treatment. In the waning scenario, the threshold drug price of ATT was <€0/year (95% CI: <€0 to €2,250) at a WTP of €20,000/QALY, and €3,250 (95% CI: <€0 to €8,000) at a WTP of €50,000/QALY. The threshold drug price was, respectively, negative in 84% and 11% of simulations, indicating that in most simulations there was no price at which ATT was cost-effective at a WTP of €20,000/QALY.

**Figure 3:**
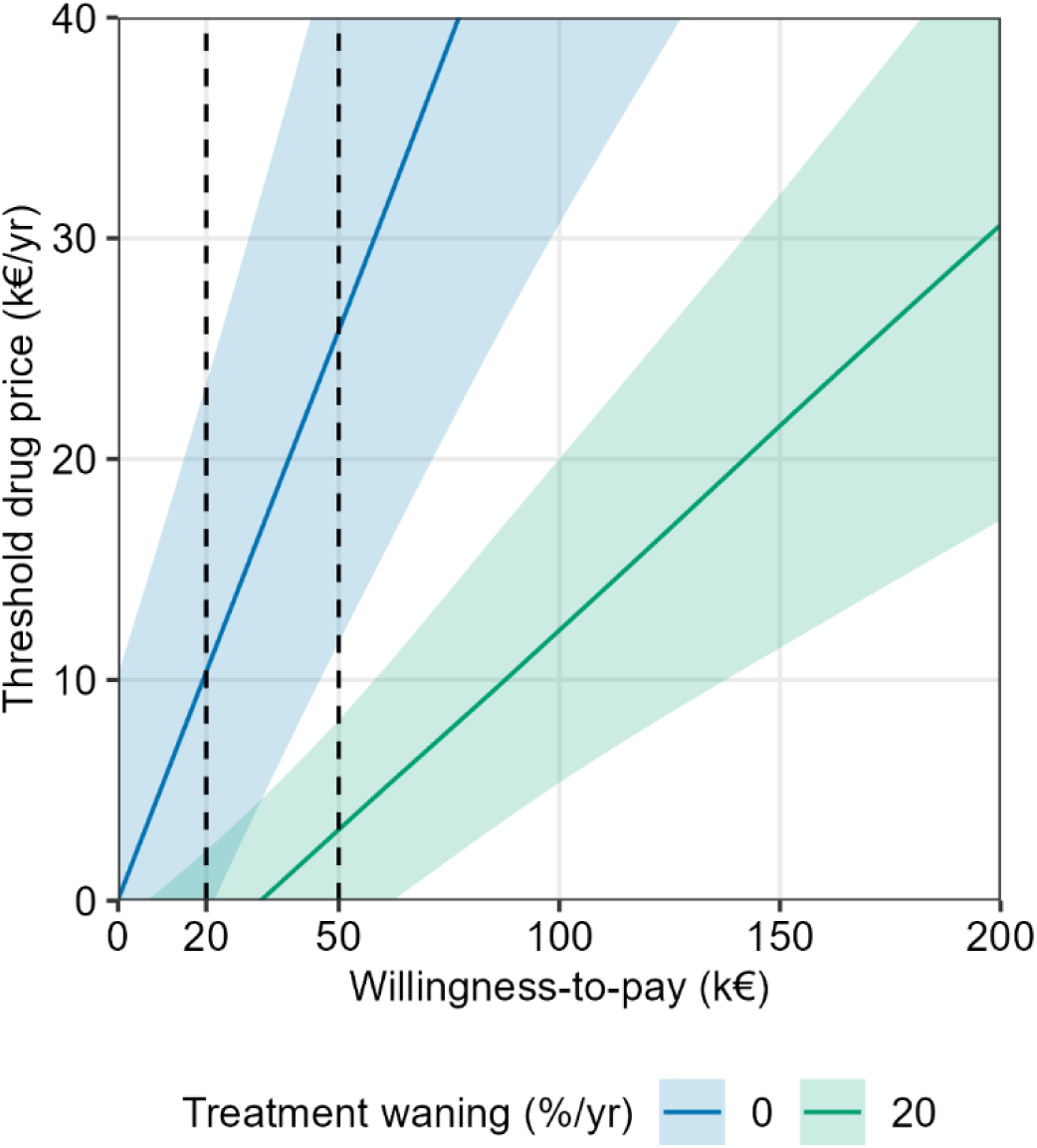
Threshold ATT prices for a range of willingness-to-pay thresholds. The lines and confidence bounds were smoothed using a “loess” function with a span of 0.5. Threshold drug prices do not include the cost related to treatment such as those of monthly infusions and monitoring MRIs. The threshold drug price of treatment could be negative if other cost changes exceeded the net monetary benefit at a given willingness to pay.

Restricting the simulated cohort to patients with MCI at baseline, the increase in QALYs in the ATT no-waning scenario was 0.74 compared to CAU and the increase in costs was €28,225. This yielded an ICER of €38,263/QALY (Figure 4). Restricting the simulated cohort to patients with mild dementia at baseline, the increase in QALYs in the ATT no-waning scenario was 0.51 compared to CAU and the increase in costs was €31,338. This yielded a substantially higher ICER of €61,814/QALY.

**Figure 4:**
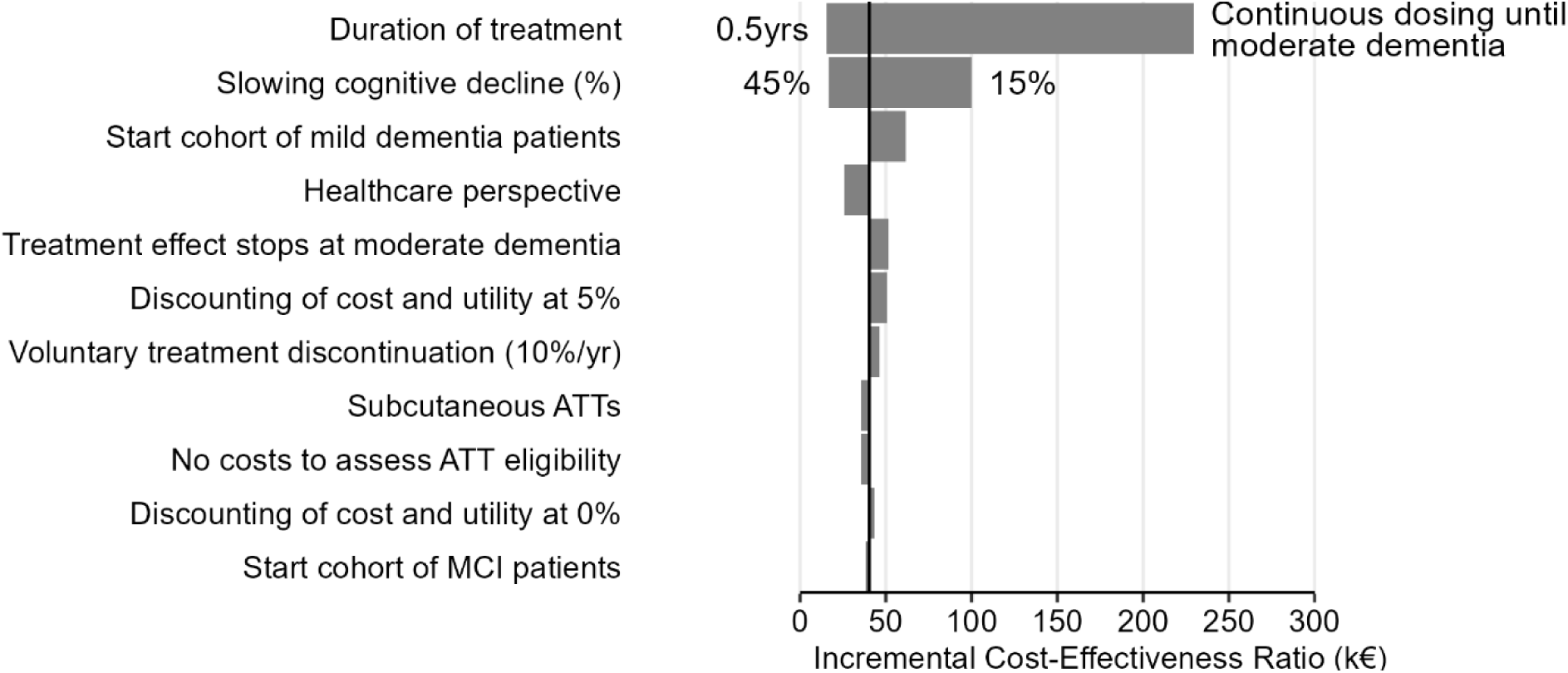
ICERs under alternative treatment, cohort, cost and discounting rate assumptions. ICERs reflect the cost in k€ per QALY gained. The dashed line indicates the no-waning base case ICER of €39,745. Abbreviations: yr= year

Parameter sensitivity analyses starting from the no-waning scenario showed that treatment duration strongly influenced the ICER (Figure 4, Supplemental Table 5). In the scenario where ATT was given until patients reached moderate dementia, the increase in mean QALY per person was 0.62 and the increase in costs was €142,269, yielding an ICER of €229,645/QALY. Changing the cost assumptions or discounting rate only slightly influenced the ICER (Supplemental Table 6).

The structural uncertainty analysis explored alternative ways of operationalising the ATT effects (Figure 5; Supplemental table 8). When ATT effects were modelled as a direct effect on functional independence rather than through cognitive decline, cost-effectiveness improved substantially in the 20% waning scenario (ICER decreasing from €151,016 to €83,484/QALY) while remaining unchanged in the no-waning scenario (€39,745/QALY vs. €40,812/QALY). When an all-or-nothing treatment effect instead of a “leaky” effect was assumed, cost-effectiveness improved in the no-waning scenario due to lower incremental costs (ICER decreasing from €39,745/QALY to €24,665/QALY). Assuming an all-or-nothing treatment effect did not change the cost-effectiveness when waning was present. Irrespective of how the ATT treatment effect was implemented in the model, effect waning had a strong effect on the cost-effectiveness estimates.

**Figure 5:**
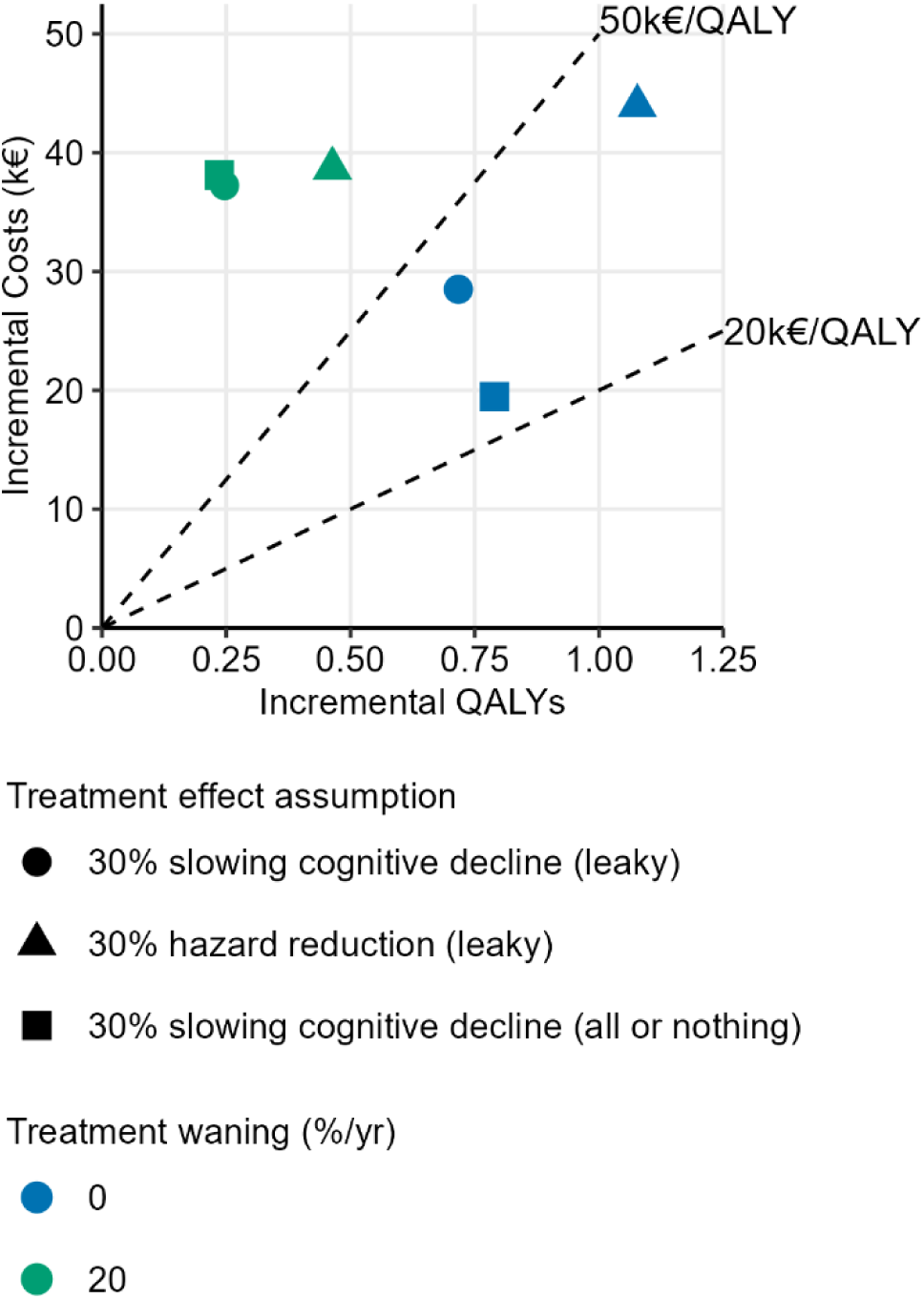
Cost and utilities of different treatment effect operationalisations. The dashed lines represent a willingness to pay threshold of €20,000/QALY and €50,000/QALY. Leaky effects indicate that all participants have the same treatment effect (30%). All-or-nothing effects indicate that 30% of the participants experience no cognitive decline and 70% of the participants decline under the care-as-usual scenario.

## Discussion

We estimated the cost-effectiveness of ATTs from a Dutch societal perspective using an AD simulation model linking cognitive decline and functional independence. If ATTs were given over eighteen months, after which treatment provision stopped and effects remained while patients were in the community (i.e. no waning), ATTs were likely to be cost-effective at a WTP of €50,000/QALY but not at €20,000/QALY. If the treatment effects waned or continuous provision of ATTs was assumed, ATTs were unlikely to be cost-effective at WTP thresholds of €20,000, €50,000, and €80,000/QALY. These results were robust to structural uncertainty in the model due to operationalisation of the treatment effects.

Two assumptions that substantially influenced the cost-effectiveness of ATTs were treatment effect waning and treatment duration. The promise of ATTs’ disease-modifying properties is that a persisting slowing of disease progression leads to an accumulation of benefits. Without a persisting disease slowing, the effect of ATTs on cognition would be similar to symptomatic treatments. Unfortunately, the randomised controlled trials could not inform us if the effects of ATTs wane over time due to their short duration. There is some evidence that clinical slowing was stable and persisting with continued lecanemab dosing compared to a historical cohort,^46, 47^ but without continued lecanemab treatment, post-trial follow-up indicated substantial treatment effect waning.^48^ Post-trial follow-up of donanemab patients showed some evidence of persisting clinical slowing after the treatment stopped compared to a historical cohort.^49^ In addition to uncertainty regarding long-term treatment effects, there is also uncertainty about the treatment duration and the corresponding costs. Based on their trial designs and the targeted amyloid isoforms, the Appropriate Use Recommendations for lecanemab advise continuous dosing,^28^ while for donanemab there is the option to stop treatment after amyloid clearance.^29^ Upcoming developments may lower treatment costs, with improved brain-barrier penetration lowering the required dosage,^50^ and subcutaneous treatment eliminating infusion costs.^51^

Our findings are broadly in line with previous cost-effectiveness studies. Like other studies, we found that treatment effect waning substantially influenced the results, though the change in cost-effectiveness we found was larger than in other studies. Our model estimated an almost five-fold increase in the ICER of ATTs in the waning scenario compared to no waning, while the other studies reported around a doubling in the ICER or threshold price.^9, 13, 16^ A possible explanation is that our model predicted the greatest utility gains in the mild dementia stage in the no-waning scenario, at which point treatment effects had already substantially decreased in the 20% waning scenario. Other studies also highlighted the importance of treatment duration for the cost-effectiveness of ATTs. Three studies investigated scenarios with limited duration of treatment provision and persisting effects.^12, 14, 16^ ATTs were only cost-effective if limited duration and persisting effects were assumed for all patients without subsequent waning.^12, 16^ Six cost-effectiveness simulation studies included continuous dosing scenarios for ATTs. Three of these studies predicted continuous dosing to be cost-effective using the AD Archimedes condition-event (ACE) microsimulation model.^17–19^ The three remaining studies with continuous dosing scenarios estimated ICERs between $258,000/QALY^11^ and €330,835/QALY^9^ with different simulation models.^16^ The studies using the AD-ACE model involved co-authors from the industry, and in contrast to other models, the AD-ACE model predicted increased clinical slowing over time due to a lower amyloid PET burden. While the range of ICERs is quite broad, the continuous dosing scenarios often led to ICERs above commonly used WTP thresholds.

Our study was novel in exploring the structural uncertainty arising from the operationalisation of ATT effects in the model. We found that the ICER decreased substantially in a scenario with 20% waning if we assumed that ATTs directly affected functional independence. Note that this type of scenario is common in other simulation models.^52^ Therefore, if our assumption holds that ATTs exert their primary effect through cognition, results from other studies are likely to be optimistic. Nonetheless, the cost-effectiveness estimates were most strongly impacted by the assumed treatment effect waning and treatment duration.

This study has two main strengths. First, our simulation model linked granular cognitive decline with loss of functional independence using statistical joint models for longitudinal and survival data. Conceptually, our model follows the common understanding of AD, where pathology leads to cognitive impairment and subsequently loss of functional independence.^20^ Because of this novel framework, the influence of structural model assumption on ATT cost-effectiveness could be evaluated. Second, we developed our simulation model using a single population of amyloid-positive MCI and dementia patients from a memory clinic. This population is highly representative of ATT-eligible patients. Other simulation models included patients without biomarker confirmed AD diagnoses to estimate part of their model.^14–19^ Due to the substantial diagnostic misclassification without amyloid biomarkers,^21^ this reduced the representativeness of these model estimates for the target population.

There are some notable limitations. First, the costs and utilities used in this study were not all sourced from the population used to develop the model, but from Dutch reference data,^33^ Swedish care data,^34^ and Western European cohorts.^35^ This may reduce the representativeness for Dutch care. Second, multiple assumptions were made to link the reported ARIA effects in trials to the monitoring and treatment cessation suggestions in the Appropriate Use Recommendations. While this might reduce the accuracy of the ARIA estimates, the overall impact of ARIA on the results was small. Third, while the entire model population was amyloid-positive, ATT-eligibility could not be ensured due to missing information on all eligibility criteria. However, the Amsterdam Dementia Cohort has a low comorbidity burden,^53^ which is a frequent reason for ATT-ineligibility,^25^ suggesting that the study population is likely to be representative of for ATT-eligible patients. Fourth, this study did not evaluate lecanemab or donanemab individually but modelled the effect of a single representative treatment to explore how treatment and modelling assumptions affected ATT cost-effectiveness. Nonetheless, the modelled treatment was informed by the lecanemab and donanemab trial. Fifth, as the ATT scenarios were compared to the current CAU, costs related to confirming AD pathology, monitoring and infusion were included in the cost of treatment as these are not part of standard clinical practice.^54^ If these improvements in the diagnostic work-up are offered to all patients with memory complaints in the future, the ICER would decrease somewhat.

To improve the accuracy of the cost-effectiveness estimates of ATTs, future studies should narrow the uncertainty surrounding highly influential parameters such as treatment effect waning. Real-world data from registries of patients on treatment combined with causal inference methodology, as well as clinical trials with longer follow-up, could provide such information.^55^ These results can then support the design of pragmatic trials that optimise treatment durations or alternative dosing schedules in order to maintain enduring treatment effects while minimising patient burden and costs.

## Conclusion

This study investigated the cost-effectiveness of ATTs in the Netherlands using an AD simulation model. We found that ATTs could be cost-effective under the assumption that treatment duration was limited to eighteen months and effects did not wane after treatment provision stopped. These results were robust to structural assumption on how ATTs affected disease progression in the model. Our results emphasise that determining the degree of the treatment effect waning in combination with treatment duration are important for an accurate assessment of the cost-effectiveness.

## Ethical approval and patient consents

The study protocol of the Amsterdam Dementia Cohort was approved by the ethical review board of the VU University Medical Centre (2016.061). Written informed consent was obtained from all patients for the use of their data for research purposes.

## Supporting information

Supplemental methods and results

Simulation scenario table

CHEER checklist

## Data Availability

Original data can be made available upon request. Simulation code is available online at: https://github.com/PieterVeere/ADC_CEA_sim

## Acknowledgements

Results in this paper used calculations by the Amsterdam UMC in project number 8632 using non-public microdata from Statistics Netherlands.

This project was funded by a Personalized Medicine Approach for Alzheimer’s Disease (ABOARD), a public-private partnership receiving funding from ZonMW (#73305095007) and Health∼Holland, Topsector Life Sciences & Health (PPP-allowance; #LSHM20106). More than <u>30 partners</u> participate in ABOARD (www.aboard-project.nl). ABOARD also receives funding from Edwin Bouw Fonds and Gieskes-Strijbisfonds. Alzheimer Center Amsterdam and Neurochemistry Laboratory Amsterdam UMC have received unrestricted funding from Alzheimer Nederland, Stichting VUmc Fonds, Genootschap tot Steun Alzheimercentrum, Alzheimer Rally, and many others. Commercial partners in consortia or for contract research: Life-MI, Brain Research Center, AVID, Winterlight labs, Nutricia, ADx Neurosciences, Roche AG, Novartis-NL, Philips, Combinostics, Danone-Nutricia, Castor, Neurocast, FujiFilm-Toyama, Quanterix, Eli Lilly, AC-Immune, Axon Neurosciences, BioConnect, Bioorchestra, Brainstorm Therapeutics, Celgene, Cognition Therapeutics, EIP Pharma, Eisai, Fujirebio, Grifols, Instant Nano Biosensors, Merck, Novo Nordisk, PeopleBio, Siemens, Vivoryon. Grant funding: NWO, ZonMW, CVON, EU-JPND, EU-IMI, EU-IHI, Alzheimer Nederland, Hersenstichting, Health∼Holland Top Sector Life Sciences & Health, Stichting Dioraphte, Gieskes Strijbis Fonds, Edwin Bouw Fonds, Pasman stichting, stichting Equilibrio, European Commission (Marie Curie International Training Network), Innovative Medicines Initiatives 3 TR, EPND, National MS Society, Alzheimer Drug Discovery Foundation, Alzheimer Association, The Selfridges Group Foundation.

Research of H.M. Broulikova was supported by the Ministry of Health of the Czech Republic, grant nr. NW24J-07-00064.

## Disclosures

P.J. van der Veere, H.M. Broulikova, E.G.B. Vijverberg, and J. Berkhof have no disclosures.

Research of **CET** is supported by the European Commission (Marie Curie International Training Network, grant agreement No 860197 (MIRIADE) and No 101119596 (TAME), Innovative Medicines Initiatives 3TR (Horizon 2020, grant no 831434) EPND (IMI 2 Joint Undertaking (JU), grant No. 101034344) and JPND (bPRIDE, CCAD), European Partnership on Metrology, co-financed from the European Union’s Horizon Europe Research and Innovation Programme and by the Participating States ((22HLT07 NEuroBioStand), Horizon Europe (PREDICTFTD, 101156175), CANTATE project funded by the Alzheimer Drug Discovery Foundation, Alzheimer Association, Michael J Fox Foundation, Health Holland, the Dutch Research Council (ZonMW), Alzheimer Drug Discovery Foundation, The Selfridges Group Foundation, Alzheimer Netherlands. CT is recipient of ABOARD, which is a public-private partnership receiving funding from ZonMW (#73305095007) and Health∼Holland, Topsector Life Sciences & Health (PPP-allowance; #LSHM20106). CT is recipient of TAP-dementia, a ZonMw funded project (#10510032120003) in the context of the Dutch National Dementia Strategy. CET has **research contracts** with Acumen, ADx Neurosciences, AC-Immune, Alamar, Aribio, Axon Neurosciences, Beckman-Coulter, BioConnect, Bioorchestra, Brainstorm Therapeutics, Celgene, Cognition Therapeutics, EIP Pharma, Eisai, Eli Lilly, Fujirebio, Instant Nano Biosensors, Novo Nordisk, Olink, PeopleBio, Quanterix, Roche, Toyama, Vivoryon. CET has **research contracts** with Acumen, ADx Neurosciences, AC-Immune, Alamar, Aribio, Axon Neurosciences, Beckman-Coulter, BioConnect, Bioorchestra, Brainstorm Therapeutics, C2N diagnostics, Celgene, Cognition Therapeutics, EIP Pharma, Eisai, Eli Lilly, Fujirebio, Instant Nano Biosensors, Merck, Muna, Novo Nordisk, Olink, PeopleBio, Quanterix, Roche, Toyama, Vaccinex, Vivoryon. She is **editor** in chief of Alzheimer Research and Therapy, and serves on editorial boards of Molecular Neurodegeneration, Alzheimer’s & Dementia, Neurology: Neuroimmunology & Neuroinflammation, Medidact Neurologie/Springer, and is committee member to define guidelines for Cognitive disturbances, and one for acute Neurology in the Netherlands. She has **consultancy/speaker contracts** for Aribio, Biogen, Beckman-Coulter, Cognition Therapeutics, Eisai, Eli Lilly, Merck, Novo Nordisk, Novartis, Olink, Roche, Sanofi and Veravas.

WF is executive director at Alzheimer Nederland, Amersfoort the Netherlands. In the past, research programs of Wiesje van der Flier have been funded by ZonMW, NWO, EU-JPND, EU-IHI, Alzheimer Nederland, Hersenstichting CardioVascular Onderzoek Nederland, Health∼Holland, Topsector Life Sciences & Health, stichting Dioraphte, Noaber foundation, Pieter Houbolt Fonds, Gieskes-Strijbis fonds, stichting Equilibrio, Edwin Bouw fonds, Pasman stichting, Philips, Biogen MA Inc, Novartis-NL, Life-MI, AVID, Roche BV, Eli-Lilly-NL, Fujifilm, Eisai, Combinostics. WF is recipient of ABOARD, which is a public-private partnership receiving funding from ZonMW (#73305095007) and Health∼Holland, Topsector Life Sciences & Health (PPP-allowance; #LSHM20106). In the past, WF has been an invited speaker at Biogen MA Inc, Danone, Eisai, WebMD Neurology (Medscape), NovoNordisk, Springer Healthcare, European Brain Council. WF has been consultant to Oxford Health Policy Forum CIC, Roche, Biogen MA Inc, Eisai, Eli-Lilly, Owkin France, Nationale Nederlanden Ventures. WF has participated in advisory boards of Biogen MA Inc, Roche, and Eli Lilly. WF has been member of the steering committee of phase 3 EVOKE/EVOKE+ studies (NovoNordisk). WF has been member of the steering committee op phase 3 Trontinemab study (Roche). All funding has been paid to Amsterdam UMC. WF was associate editor of Alzheimer, Research & Therapy in 2020/2021. WF was associate editor at Brain 2021-2025.

WF is chair of the Scientific Leadership Group of InRAD.

WF is member of Supervisory Board (Raad van Toezicht) Trimbos Instituut.

L.E. Collij has received research support from GE Healthcare, Life Molecular Imaging, and Springer Healthcare (funded by Eli Lilly), paid to their institution.

R. Handels received outside this study consulting fees in the past 36 months from Lilly Nederland and from the Institute for Medical Technology Assessment (paid to institution).

## References

1. Ferrari AJ, Santomauro DF, Aali A, Abate YH, Abbafati C, Abbastabar H, et al. Global incidence, prevalence, years lived with disability (YLDs), disability-adjusted life-years (DALYs), and healthy life expectancy (HALE) for 371 diseases and injuries in 204 countries and territories and 811 subnational locations, 1990-2021: a systematic analysis for the Global Burden of Disease Study 2021. The Lancet. 2024;403(10440):2133–61.

2. Velandia PP, Miller-Petrie MK, Chen C, Chakrabarti S, Chapin A, Hay S, et al. Global and regional spending on dementia care from 2000-2019 and expected future health spending scenarios from 2020-2050: An economic modelling exercise. EClinicalMedicine. 2022;45:101337.

3 World Health Organisation. Ageing and health 2024 [Available from: https://www.who.int/news-room/fact-sheets/detail/ageing-and-health.

4. Xu H, Garcia-Ptacek S, Jönsson L, Wimo A, Nordström P, Eriksdotter M. Long-term Effects of Cholinesterase Inhibitors on Cognitive Decline and Mortality. Neurology. 2021;96(17):e2220–e30.

5. Sims JR, Zimmer JA, Evans CD, Lu M, Ardayfio P, Sparks J, et al. Donanemab in Early Symptomatic Alzheimer Disease: The TRAILBLAZER-ALZ 2 Randomized Clinical Trial. JAMA. 2023;330(6):512–27.

6. van Dyck CH, Swanson CJ, Aisen P, Bateman RJ, Chen C, Gee M, et al. Lecanemab in Early Alzheimer’s Disease. N Engl J Med. 2023;388(1):9–21.

7 Eisai. EISAI’S APPROACH TO U.S. PRICING FOR LEQEMBI™ (LECANEMAB), A TREATMENT FOR EARLY ALZHEIMER’S DISEASE, SETS FORTH OUR CONCEPT OF “SOCIETAL VALUE OF MEDICINE” IN RELATION TO “PRICE OF MEDICINE” 2023 [Available from: https://www.eisai.com/news/2023/news202302.html.

8 Lilly. Lilly’s Kisunla™ (donanemab-azbt) Approved by the FDA for the Treatment of Early Symptomatic Alzheimer’s Disease 2024 [Available from: https://investor.lilly.com/news-releases/news-release-details/lillys-kisunlatm-donanemab-azbt-approved-fda-treatment-early.

9. Wimo A, Handels R, Blennow K, Kirsebom B-E, Selnes P, Bon J, et al. Cost-effectiveness of diagnosing and treating patients with early Alzheimer’s disease with anti-amyloid treatment in a clinical setting. Journal of Alzheimer’s Disease. 2025;104(4):1167–84.

10. Wright AC, Lin GA, Whittington MD, Agboola F, Herron-Smith S, Rind D, et al. The effectiveness and value of lecanemab for early Alzheimer disease: A summary from the Institute for Clinical and Economic Review’s California Technology Assessment Forum. Journal of Managed Care & Specialty Pharmacy. 2023;29(9):1078–83.

11 Lin GA, Whittington MD, Wright A, Agboola F, Herron-Smith S, Pearson SD, et al. Beta-Amyloid Antibodies for Early Alzheimer’s Disease: Effectiveness and Value; Evidence Report. Institute for Clinical and Economic Review,. 2023.

12. Boustani M, Doty EG, Garrison LP, Jr., Smolen LJ, Klein TM, Murphy DR, et al. Estimating the Economically Justifiable Price of Limited-Duration Treatment with Donanemab for Early Symptomatic Alzheimer’s Disease in the United States. Neurol Ther. 2024;13(6):1641–59.

13. Xia X, Aye S, Frisell O, Aho E, Handels R, Li Y, et al. The Cost-Effective Price of Lecanemab for Patients with Early Alzheimer’s Disease in Sweden. Pharmacoeconomics. 2025.

14. Ross EL, Weinberg MS, Arnold SE. Cost-effectiveness of Aducanumab and Donanemab for Early Alzheimer Disease in the US. JAMA Neurol. 2022;79(5):478–87.

15. Shin S, Kim M, Hong SH. Economic Evaluation of Lecanemab for Early Symptomatic Alzheimer’s Disease in South Korea. PharmacoEconomics - Open. 2025;9(5):793–804.

16. Handels R, Herring WL, Grimm S, Sköldunger A, Winblad B, Wimo A, et al. New IPECAD Open-Source Model Framework for the Health Technology Assessment of Early Alzheimer’s Disease Treatment: Development and Use Cases. Value in Health. 2025;28(4):511–8.

17. Igarashi A, Azuma MK, Zhang Q, Ye W, Sardesai A, Folse H, et al. Predicting the Societal Value of Lecanemab in Early Alzheimer’s Disease in Japan: A Patient-Level Simulation. Neurol Ther. 2023;12(4):1133–57.

18. Tahami Monfared AA, Ye W, Sardesai A, Folse H, Chavan A, Kang K, et al. Estimated Societal Value of Lecanemab in Patients with Early Alzheimer’s Disease Using Simulation Modeling. Neurol Ther. 2023;12(3):795–814.

19. Tahami Monfared AA, Tafazzoli A, Chavan A, Ye W, Zhang Q. The Potential Economic Value of Lecanemab in Patients with Early Alzheimer’s Disease Using Simulation Modeling. Neurol Ther. 2022;11(3):1285–307.

20. Liu-Seifert H, Siemers E, Price K, Han B, Selzler KJ, Henley D, et al. Cognitive Impairment Precedes and Predicts Functional Impairment in Mild Alzheimer’s Disease. J Alzheimers Dis. 2015;47(1):205–14.

21. Beach TG, Monsell SE, Phillips LE, Kukull W. Accuracy of the clinical diagnosis of Alzheimer disease at National Institute on Aging Alzheimer Disease Centers, 2005-2010. J Neuropathol Exp Neurol. 2012;71(4):266–73.

22. Van der Veere PJ, Broulikova HM, Hoogland J, Handels R, Vijverberg EGB, Van Harten AC, et al. Developing a microsimulation model of Alzheimer’s Disease for the evaluation of disease modifying therapies. Under Review.

23. Hickey GL, Philipson P, Jorgensen A, Kolamunnage-Dona R. Joint modelling of time-to-event and multivariate longitudinal outcomes: recent developments and issues. BMC medical research methodology. 2016;16:1–15.

24. Perneczky R, Wagenpfeil S, Komossa K, Grimmer T, Diehl J, Kurz A. Mapping Scores Onto Stages: Mini-Mental State Examination and Clinical Dementia Rating. The American Journal of Geriatric Psychiatry. 2006;14(2):139–44.

25. Claus JJ, Vom Hofe I, van Ijlzinga Veenstra A, Licher S, Seelaar H, de Jong FJ, et al. Generalizability of trial criteria on amyloid-lowering therapy against Alzheimer’s disease to individuals with mild cognitive impairment or early Alzheimer’s disease in the general population. Eur J Epidemiol. 2025;40(3):327–37.

26. van der Flier WM, Scheltens P. Amsterdam Dementia Cohort: Performing Research to Optimize Care. J Alzheimers Dis. 2018;62(3):1091–111.

27. Greenberg SM, Bax F, van Veluw SJ. Amyloid-related imaging abnormalities: manifestations, metrics and mechanisms. Nature Reviews Neurology. 2025;21(4):193–203.

28. Cummings J, Apostolova L, Rabinovici GD, Atri A, Aisen P, Greenberg S, et al. Lecanemab: Appropriate Use Recommendations. J Prev Alzheimers Dis. 2023;10(3):362–77.

29. Rabinovici GD, Selkoe DJ, Schindler SE, Aisen P, Apostolova LG, Atri A, et al. Donanemab: Appropriate use recommendations. The Journal of Prevention of Alzheimer’s Disease. 2025;12(5):100150.

30. Landeiro F, Mughal S, Walsh K, Nye E, Morton J, Williams H, et al. Health-related quality of life in people with predementia Alzheimer’s disease, mild cognitive impairment or dementia measured with preference-based instruments: a systematic literature review. Alzheimer’s Research & Therapy. 2020;12(1):154.

31. Luengo-Fernandez R, Gray AM, Bull L, Welch S, Cuthbertson F, Rothwell PM. Quality of life after TIA and stroke: ten-year results of the Oxford Vascular Study. Neurology. 2013;81(18):1588–95.

32 Alzheimer-Medikament Leqembi kommt ab September auf den Markt. Die Zeit. 2025 2025–08–31.

33. Zorginstituut Nederland. Verdiepingsmodule Kostenhandleiding. 2024.

34. Aye S, Frisell O, Zetterberg H, Skillbäck TB, Kern S, Eriksdotter M, et al. Costs of Care in Relation to Alzheimer’s Disease Severity in Sweden: A National Registry-Based Cohort Study. PharmacoEconomics. 2025;43(2):153–69.

35. Handels R, Hataiyusuk S, Wimo A, Sköldunger A, Bakker C, Bieber A, et al. Informal care for people with dementia in Europe. The Journal of Prevention of Alzheimer’s Disease. 2025;12(1):100015.

36 Centraal Bureau voor de Statistiek. Consumentenprijzen; prijsindex 2025 [Available from: https://www.cbs.nl/nl-nl/cijfers/detail/83131NED.

37. Zorginstituut Nederland. Richtlijn voor het uitvoeren van economische evaluaties in de gezondheidszorg. 2024.

38. Livingston G, Huntley J, Liu KY, Costafreda SG, Selbæk G, Alladi S, et al. Dementia prevention, intervention, and care: 2024 report of the Lancet standing Commission. The Lancet. 2024;404(10452):572–628.

39. Enzing J, Knies S, Zwaap J. Beoordelingskader kosteneffectiviteit van zorg. In: Zorginstituut Nederland, editor. Diemen2024.

40. Polder J, Van Exel J, Kahlman J, Knies S, Mierau J, Vermeulen W, et al. Preventie op waarde schatten. Advies technische werkgroep kosten en baten van preventie: ZonMW; 2023.

41. Husereau D, Drummond M, Augustovski F, de Bekker-Grob E, Briggs AH, Carswell C, et al. Consolidated Health Economic Evaluation Reporting Standards 2022 (CHEERS 2022) Statement: Updated Reporting Guidance for Health Economic Evaluations. Value Health. 2022;25(1):3–9.

42. Team RC. R: A Language and Environment for Statistical Computing. 4.4.3 ed: R Foundation for Statistical Computing; 2025.

43 Methylprednisolon: Farmacotherapeutisch Kompas; [Available from: https://www.farmacotherapeutischkompas.nl/bladeren/preparaatteksten/m/methylprednisolon.

44. Hamilton E, Van Boxmeer L. Kosten in Kaart - Neurologie: Vereniging Arts-Assistenten in opleiding tot Neuroloog; 2022 [

45 Hamer HM. APOE gevonden genmutaties [identificatie] in bloed of weefsel m.b.v.moleculair genetische methode nkvc.nl: Nederlandse Vereniging voor Klinische Chemie en Laboratoriumgeneeskunde; [Available from: https://www.nvkc.nl/professional/wie-doet-wat-database/216192-a003-ba19cd9d-c13e412a92ce8dd82e07a76e.

46. van Dyck C. Does the Current Evidence Base Support Lecanemab Continued Dosing for Early Alzheimer’s Disease? Alzheimer’s Association International Conference; Philadelphia 2024.

47 Early Alzheimer’s Patients Continue to Benefit from Four Years of LEQEMBI® (lecanemab-irmb) Therapy New Clinical Data Presented at AAIC [press release]. Tokyo and Cambridge, Massachusetts, 2025–07–31 2025.

48. McDade E, Cummings JL, Dhadda S, Swanson CJ, Reyderman L, Kanekiyo M, et al. Lecanemab in patients with early Alzheimer’s disease: detailed results on biomarker, cognitive, and clinical effects from the randomized and open-label extension of the phase 2 proof-of-concept study. Alzheimer’s Research & Therapy. 2022;14(1):191.

49. Zimmer JA, Sims JR, Evans CD, Nery ESM, Wang H, Wessels AM, et al. Donanemab in early symptomatic Alzheimer’s disease: results from the TRAILBLAZER-ALZ 2 long-term extension. The Journal of Prevention of Alzheimer’s Disease. 2025:100446.

50. Grimm HP, Schumacher V, Schäfer M, Imhof-Jung S, Freskgård PO, Brady K, et al. Delivery of the Brainshuttle™ amyloid-beta antibody fusion trontinemab to non-human primate brain and projected efficacious dose regimens in humans. MAbs. 2023;15(1):2261509.

51 FDA Approves LEQEMBI® IQLIK™ (lecanemab-irmb) Subcutaneous Injection for Maintenance Dosing for the Treatment of Early Alzheimer’s Disease [press release]. Tokyo and Cambridge Massachusetts, 2025–08–29 2025.

52. Handels R, Herring WL, Kamgar F, Aye S, Tate A, Green C, et al. IPECAD Modeling Workshop 2023 Cross-Comparison Challenge on Cost-Effectiveness Models in Alzheimer’s Disease. Value Health. 2024;28(4):497–510.

53. van der Veere PJ, Hoogland J, Visser LN, Van Harten AC, Rhodius-Meester HF, Sikkes SA, et al. Predicting Cognitive Decline in Amyloid-Positive Patients With Mild Cognitive Impairment or Mild Dementia. Neurology. 2024;103(3):e209605.

54. Dementie; 2.7: Liquoronderzoek dementie. In: Richtlijnendatabase FMS, editor. 2021.

55. Perneczky R, Darby D, Frisoni GB, Hyde R, Iwatsubo T, Mummery CJ, et al. Real-world datasets for the International Registry for Alzheimer’s Disease and Other Dementias (InRAD) and other registries: An international consensus. The Journal of Prevention of Alzheimer’s Disease. 2025;12(4):100096.

