## Supplemental methods and results for "Cost-effectiveness of amyloid-targeting therapies in a memory clinic population; a simulation study"

Supplementary methods

*ARIA probability*

We calculated the probability of developing ARIA, the distribution of ARIA MRI severity, and the proportion of ARIA that are symptomatic. Data from APOE ε4 heterozygotes and non-carrier were combined from the donanemab and lecanemab trials if both reported it. Note that the probability of ARIA is lower for lecanemab than for donanemab,^1^ and that donanemab has a lower ARIA probability with longer titration.^2^ Where data from only one drug were available, they were assumed to be applicable for all ATTs.

Due to limited data availability, four further assumptions were made: (1) ARIA-H and ARIA-E had the same probability to give symptoms; (2) the probability of becoming symptomatic was equal across MRI severities of ARIA-A and ARIA-H; (3) the probability of ARIA occurring was uniformly distributed over the trial duration; (4) deaths were assumed to occur only for radiologically severe, symptomatic ARIA cases.

Based on the combined ARIA counts from ε4 heterozygotes and non-carriers of the donanemab and lecanemab trials, the probability of developing ARIA was 29% over an eighteen-month period.^1^ Supplemental table 2 outlines the distribution of ARIA MRI severity, the probability of an ARIA occurrence being symptomatic, and the probability that radiologically severe, symptomatic aria was deadly. Supplemental table 3 shows the annual probability of ARIA by MRI severity.

*ARIA costs*

The cost of MRI monitoring was set to the sum of costs of the MRI scan and the consultation with a neurologist (supplemental table 3). The cost for treatment of symptomatic severe aria is shown in supplemental table 4.

*Informal care hours*

Care partners of patients included in the Amsterdam Dementia Cohort were asked to complete the Resource Utilisation in Dementia (RUD) Lite questionnaire since 2021.^3^ The mean hours of informal care per day was calculated using data on 44 patients with amyloid-positive mild cognitive impairment from the Amsterdam Dementia Cohort for whom care partners completed the RUD Lite within two years of the diagnosis. The number of informal care hours was the sum of hours per day spent on assisting activities of daily living, assisting in spent on instrumental activities of daily living, and on supervision of the patient, divided by the proportion of informal care that was performed by the care partner filling in the RUD Lite.

*Statistical joint models and Weibull models (model 1 to 4)*

The microsimulation model was developed in a prior study and was built from four statistical models that describe transitions from MCI to living with dementia in the community, from dementia in the community to institutionalisation or death, and from institutionalisation to death. The core of the microsimulation model was formed by joint models, which combine longitudinal markers and a time-to-event model.^4^ The longitudinal marker in this study was the Mini-Mental State examination (MMSE), a measure of cognition, modelled as a quadratic function over time using linear mixed models. The modelled MMSE values were included as time-varying predictors in the time-to-event part of the joint models, modifying the risk of transitioning to the next state.

Model 1 describes the transition from MCI to dementia using a joint model with endpoints MMSE and time to dementia (represented in formula (1) and (2)). The hazard of the time-to-event model was described by a piecewise constant spline function. Model 2 describes the transition from dementia to institutionalisation or death outside an institution using a joint model with endpoints MMSE, time to institutionalisation and time to death (model 2; formulas (3-5)). The two time-to-event endpoints were described by a competing risk model and the hazards were described by piecewise constant spline functions. Model 2 was fitted to data of the first four years of follow-up of participants with dementia at baseline, because sufficient MMSE data was only available for the first four years of follow-up. Model 3 describes the transition from dementia to institutionalisation or death outside an institution from four years of follow-up onwards using a Weibull competing risk survival model. Model 3 was fitted to data of participants who were alive and not institutionalised four years after their dementia diagnosis. Model 4 describes the transition from institution to death using a Weibull model.

In all statistical models, included variables were selected based on the literature. Age and sex were included in all models. For the MMSE trajectory in the MCI stage (model 1), variables were included on cerebrospinal fluid (CSF) concentration of Aβ42, CSF concentration of pTau-181 and the magnetic resonance imaging medial temporal lobe atrophy score. In the time-to-event portion of model 2, and model 3 and 4, a variable indicating whether a patient lived together with a care partner at baseline was included.^5^ The predicted MMSE at four years by model 2 was included as a variable in model 3.

The equations for model 1 are:

$${Mᵢ}^{(1)}(t) = {\beta₀}^{(1)} +{\beta₁}^{(1)}\cdot t + {\beta₂}^{(1)}\cdot t^{2} + {\beta₃}^{(1)}\cdot Ageᵢ + {\beta₄}^{(1)}\cdot Sexᵢ + {\beta₅}^{(1)}\cdot ABetaᵢ + {\beta₆}^{(1)}\cdot pTauᵢ + {\beta₇}^{(1)}\cdot MTAᵢ + {\beta₈}^{(1)}\cdot Ageᵢ\cdot t + {\beta₉}^{(1)}\cdot Sexᵢ\cdot t + {\beta₁₀}^{(1)}\cdot ABetaᵢ\cdot t + {\beta₁₁}^{(1)}\cdot pTauᵢ\cdot t + {\beta₁₂}^{(1)}\cdot MTAᵢ\cdot t + {b₀ᵢ}^{(1)} + {b₁ᵢ}^{(1)}\cdot t + {\varepsilonᵢ}^{(1)}(t)$$

(1)

where:

- ${Mᵢ}^{(1)}(t)$: MMSE in model 1 for subject *i* at time *t*

${\beta₀}^{(1)}$ to ${\beta₁₂}^{(1)}$: fixed effects model 1

- $t$: time since MCI diagnosis in years
- ${b₀ᵢ}^{(1)}$ and ${b₁ᵢ}^{(1)}$: subject specific normally distributed random intercepts and slopes model 1
- ${\varepsilonᵢ}^{(1)}\boldsymbol{\sim}N(0,\sigma^{2})$: residual error longitudinal marker model 1.

and

$${hᵢ}^{(1)}(t) = {h₀}^{(1)}(t)\cdot exp(\alpha^{(1)}\cdot{mᵢ}^{(1)}(t) + {\gamma₁}^{(1)}\cdot Ageᵢ + {\gamma₂}^{(1)}\cdot Sexᵢ)$$

(2)

where:

- ${hᵢ}^{(1)}(t)$: the hazard in model 1 for subject *i* at time *t*
- ${h₀}^{(1)}(t)$: the baseline hazard function modelled using splines in model 1
- $\alpha^{(1)}$: the association coefficient linking MMSE to the hazard in model 1
- ${mᵢ}^{(1)}(t)$: the latent MMSE (ignoring the error term) for subject *i* at moment *t* from the longitudinal marker part of model 1
- ${\gamma₁}^{(1)}$ and ${\gamma₂}^{(1)}$: the coefficients of age and sex.

The equations of model 2 are

$${Mᵢ}^{(2)}(t) = {\beta₀}^{(2)} +{\beta₁}^{(2)}\cdot t + {\beta₂}^{(2)}\cdot t^{2} + {\beta₃}^{(2)}\cdot Ageᵢ + {\beta₄}^{(2)}\cdot Sexᵢ + {\beta₅}^{(2)}\cdot Ageᵢ\cdot t + {\beta₆}^{(2)}\cdot Sexᵢ\cdot t + {b₀ᵢ}^{(2)} + {b₁ᵢ}^{(2)}\cdot t + {\varepsilonᵢ}^{(2)}(t)$$

(3)

where:

- ${Mᵢ}^{(2)}(t)$: MMSE in model 2 for subject *i* at time *t*
- ${\beta₀}^{(2)}$ to ${\beta_{6}}^{(2)}$: fixed effects in model 2
- $t$: time since dementia diagnosis in years
- ${b₀ᵢ}^{(2)}$ and ${b₁ᵢ}^{(2)}$: subject specific normally distributed random intercepts and slopes model 2
- ${\varepsilonᵢ}^{(2)}\boldsymbol{\sim}N(0,\sigma^{2})$: residual error longitudinal marker model 2.

and

$$hᵢ^{1;(2)}\left( t \right)= {h₀}^{1;(2)}(t)\cdot\exp({\alpha₁}^{(2)}\cdot{mᵢ}^{(2)}(t) + {\gamma₁}^{(2)}\cdot Ageᵢ + {\gamma₂}^{(2)}\cdot Sexᵢ + {\gamma₃}^{(2)}\cdot Carepartnerᵢ)$$

(4)

$${hᵢ}^{2;(2)}(t) = {h₀}^{2;(2)}(t)\cdot\exp({\alpha₂}^{(2)}\cdot{mᵢ}^{(2)}(t) + {\gamma₁}^{(2)}\cdot Ageᵢ + {\gamma₂}^{(2)}\cdot Sexᵢ + {\gamma₃}^{(2)}\cdot Carepartnerᵢ)$$

(5)

where:

- $hᵢ^{1;(2)}(t)$ and $hᵢ^{2;(2)}(t)$: the cause-specific hazards of institutionalisation and death in model 2
- ${h₀}^{1;(2)}(t)$ and ${h₀}^{2;(2)}(t)$: the cause-specific baseline hazard modelled using splines in model 2
- ${\alpha₁}^{(2)}$ and ${\alpha₂}^{(2)}$: the association coefficients of MMSE, specific for time-to-event endpoint in model 2
- ${mᵢ}^{(2)}(t)$: the latent MMSE (ignoring the error term) for subject *i* at moment *t* from the longitudinal marker part of model 2
- ${\gamma₁}^{(2)}$ to ${\gamma₃}^{(2)}$: The shared coefficients of age, sex, and living with a care partner in model 2.

*Structural uncertainty analysis*

The statistical joint models were replaced by Weibull hazard models except the transition from MCI to death, which was taken from Dutch life tables. The transition from MCI to dementia was estimated using patients with MCI at baseline (model 1; supplemental table 8). The transitions from dementia to the competing events of institutionalisation and death outside of an institution were estimated using patients with dementia at baseline and those who converted from MCI to dementia (model 2; supplemental table 9). Time was set to zero for the MCI patients at the conversion time and their age was updated based on their time in the MCI state. The transition from institutionalisation to death was estimated using all patients with a registered institutionalisation (model 3; supplemental table 10), with time set to zero at institutionalisation. Age and sex were included as covariates in all models. In model 2, covariates were also added indicating if a patient lived with a care partner and if a patient had previously converted from MCI to dementia. In model 3, a covariate was added indicating if a patient lived with a care partner prior to institutionalisation.

In the simulations, time spent in the MCI, dementia and institutionalised states was extracted from the models using inverse transform sampling. The age of the simulated patient was updated after each transition. To determine the time in the mild, moderate and severe dementia states for the application of costs and utilities, MMSE values were simulated from the estimated MMSE component of the joint models in parallel with simulated time-to-event values from the Weibull hazard models. This was done to keep the costs and utilities in the dementia state comparable between the joint modelling and Weibull modelling approaches.

In the ATT scenarios, the treatment effect was implemented as an instantaneous 30% reduction of the hazard of leaving the MCI and the dementia in the community states. The MMSE trajectory was also assumed to slow by 30%. The treatment effect in the dementia state stopped when an MMSE of 20 (moderate dementia) was reached.

*Derivation of variance around slowing of cognitive decline*

The variance around the 30% slowing of cognitive decline in the model was based on the variance around the treatment effect on the primary endpoint in the phase 3 randomised controlled trials of lecanemab and donanemab, which was the Clinical Dementia Rating Scale Sum of Boxes.^6, 7^ In the donanemab trial, the percentage slowing was 28.9% (95% CI: 18.26% to 39.53%). For lecanemab, the percentage slowing was approximated by dividing the difference in change from baseline after eighteen months between the treatment group and placebo group (-0.45 [95% CI: -0.67 to -0.23]) by change from baseline after eighteen months in the placebo group (1.66), resulting in a percentage slowing of 27.1% (95% CI: 13.9% to 40.4%). The pooled standard error of the slowing in decline due to donanemab (SE: 5.4%) and lecanemab (SE: 6.8%) was 4.2%.

Supplementary results

The threshold price of ATT for the no-waning scenario compared to CAU was €41,500/year (95% CI: €23,500 to €66,250) at a WTP of €80,000/QALY. The threshold price of the ATT 20% annual waning scenario compared to CAU was €8,500 (95% CI: €2,500 to €15,250) at a WTP of €80,000/QALY.

References

1.	Greenberg SM, Bax F, van Veluw SJ. Amyloid-related imaging abnormalities: manifestations, metrics and mechanisms. Nature Reviews Neurology. 2025;21(4):193–203.

2.	Wang H, Serap Monkul Nery E, Ardayfio P, Khanna R, Otero Svaldi D, Gueorguieva I, et al. Modified titration of donanemab reduces ARIA risk and maintains amyloid reduction. Alzheimers Dement. 2025;21(4):e70062.

3.	Wimo A, Winblad B. Resource utilisation in dementia: RUD Lite. Brain Aging. 2003;3(1):48–59.

4.	Hickey GL, Philipson P, Jorgensen A, Kolamunnage-Dona R. Joint modelling of time-to-event and multivariate longitudinal outcomes: recent developments and issues. BMC medical research methodology. 2016;16:1–15.

5.	Belger M, Haro JM, Reed C, Happich M, Argimon JM, Bruno G, et al. Determinants of time to institutionalisation and related healthcare and societal costs in a community-based cohort of patients with Alzheimer’s disease dementia. The European Journal of Health Economics. 2019;20:343–55.

6.	Sims JR, Zimmer JA, Evans CD, Lu M, Ardayfio P, Sparks J, et al. Donanemab in Early Symptomatic Alzheimer Disease: The TRAILBLAZER-ALZ 2 Randomized Clinical Trial. JAMA. 2023;330(6):512–27.

7.	van Dyck CH, Swanson CJ, Aisen P, Bateman RJ, Chen C, Gee M, et al. Lecanemab in Early Alzheimer's Disease. N Engl J Med. 2023;388(1):9–21.

8.	Zorginstituut Nederland. Verdiepingsmodule Kostenhandleiding. 2024.

9.	Hamilton E, Van Boxmeer L. Kosten in Kaart - Neurologie: Vereniging Arts-Assistenten in opleiding tot Neuroloog; 2022 [

10.	Hamer HM. APOE gevonden genmutaties [identificatie] in bloed of weefsel m.b.v.moleculair genetische methode nkvc.nl: Nederlandse Vereniging voor Klinische Chemie en Laboratoriumgeneeskunde; [Available from: <https://www.nvkc.nl/professional/wie-doet-wat-database/216192-a003-ba19cd9d-c13e412a92ce8dd82e07a76e>.

11.	Methylprednisolon: Farmacotherapeutisch Kompas; [Available from: <https://www.farmacotherapeutischkompas.nl/bladeren/preparaatteksten/m/methylprednisolon>.

12.	Aye S, Frisell O, Zetterberg H, Skillbäck TB, Kern S, Eriksdotter M, et al. Costs of Care in Relation to Alzheimer’s Disease Severity in Sweden: A National Registry-Based Cohort Study. PharmacoEconomics. 2025;43(2):153–69.

13.	Handels R, Hataiyusuk S, Wimo A, Sköldunger A, Bakker C, Bieber A, et al. Informal care for people with dementia in Europe. The Journal of Prevention of Alzheimer's Disease. 2025;12(1):100015.

14.	Wu H, Godfrey AJR. R: The Weibull Distribution [Available from: <https://search.r-project.org/CRAN/refmans/ExtDist/html/Weibull.html>.

15.	Van der Veere PJ, Broulikova HM, Hoogland J, Handels R, Vijverberg EGB, Van Harten AC, et al. Developing a microsimulation model of Alzheimer’s Disease for the evaluation of disease modifying therapies. Under Review.

**Supplemental Table 1**: MRI severity stage distribution, symptomatic proportion of ARIA, and proportion deadly ARIA

|  | Proportion per severity stage^6^ | Proportion symptomatic^1^* | | Percentage of symptomatic cases that were deadly |
| --- | --- | --- | --- | --- |
|  |  | Asymptomatic | Symptomatic |  |
| Mild | 58% (195/337) | 78% (655/845) | 22% (190/845) | 0%† |
| Moderate | 23% (78/337) | 78% (655/845) | 22% (190/845) | 0%† |
| Severe | 19% (64/337) | 78% (655/845) | 22% (190/845) | 14%‡ |

**To derive the percentage of symptomatic ARIA, the number of people with symptomatic ARIA in the lecanemab and donanemab trials (190; 63 + 127) was divided by the total number of people with ARIA in the lecanemab and donanemab trials (n= 845; 63 + 211 + 127 + 444). The percentage of asymptomatic ARIA is one minus the percentage of symptomatic ARIA. All numbers are derived from table 1 of Greenberg, Bax and Van Veluw (2025)^1^*

*†Assumed*

*‡ The total number of ARIA in the treatment arm of the phase 3 lecanemab trial in the APOE e4 negative and heterozygote groups was: 33 + 67 + 52 + 15 = 167.^7^ The total number of ARIA in the treatment arm of the phase 3 donanemab trial in the APOE e4 negative and heterozygote groups was: 40 + 103 + 48 + 146 = 337.^6^ Of these 504 ARIA (337 + 157), 504 * 0.19 = 96 would have been severe, 96 * 22 of which would have been symptomatic. There were three deadly ARIA’s over the treatment arms of both trial and we assume ARIA deaths only occur in the severe symptomatic ARIA cases, thus 3/22 = 14% were deadly.*

**Supplemental Table 2**: Yearly ARIA probability by MRI severity.

| MRI severity | Yearly probability of ARIA |
| --- | --- |
| Total | 20%* |
| Mild | 11% |
| Moderate | 5% |
| Severe | 4% |

** Based on table 2 of Greenberg, Bax and Van Veluw, the number of ARIA-E and ARIA-H in the APOE e4 negatives and heterozygotes divided by the total APOE e4 negatives and heterozygotes*

**Supplemental Table 3**: Costs of dementia diagnosis

|  | Cost | Source |
| --- | --- | --- |
| **General memory clinic work-up** |  |  |
| Out-patient visit neurologist | 120 | ^8^ |
| Neuropsychological testing | 100,67 | Inhouse data |
| Multidisciplinary meeting to make diagnosis* | 48 | ^8^ |
| MRI | 254 | ^8^ |
| **Additional work-up for ATT eligibility** |  |  |
| Lumbar punction and assay of AB1-42 and pTau-181 | 307,08 | ^9^ |
| APOE genotyping | 234,84 | ^10^ |

** Calculated by assuming three medical specialist would spend on average ten minutes discussing a case, using the hourly cost of a medical specialist provided in the ZiN costing manual.*^8^

*Prices are in 2022 euros*

**Supplemental Table 4**: Costs of symptomatic severe ARIA

|  | Cost | Source |
| --- | --- | --- |
| Hospitalisation for five days | 644 | ^8^ |
| Methylpredinsolon, 5 grams | 150 | ^11^ |
| Out-patient visit with neurologist | 120 | ^8^ |

*Estimates rounded to the nearest euro are presented. Prices are in 2022 euros.*

**Supplemental Table 5**: Costs per year in each state

|  | Informal care (hours * price * 365) | Standard error | Formal care | Standard error | Sources |
| --- | --- | --- | --- | --- | --- |
| Mild cognitive impairment | 6862 (0.91 * 18.80 * 365) | 2127 (0.31 * 18.80 * 365) | 6135 | 1062 | ^12^, own data |
| Mild community dementia | 18966 (2.76 * 18.80 * 365) | 4080 (0.59 * 18.80 * 365) | 12011 | 1211 | ^12, 13^ |
| Moderate community dementia | 25797 (3.76 * 18.80 * 365) | 4433 (0.65 * 18.80 * 365) | 20210 | 2494 | ^12, 13^ |
| Severe community dementia | 35177 (5.13 * 18.80 * 365) | 4157 (0.61 * 18.80 * 365) | 42009 | 11113 | ^12, 13^ |
| Institutionalisation |  |  | 110688 (290 * 365) + 4838 | NA | ^8, 12^ |

*Estimates rounded to the nearest euro are presented. Price per hour of informal care and institutional care per day are sourced from the ZiN costing manual.^8^ Hours of informal care were sources from in-house data in the MCI state and the publication of Handels et al. (2025) for the dementia states.*^13^ *Prices are in 2022 euros.*

**Supplemental Table 5:** ICERs for the scenarios after changing the treatment effect

| Scenario | CAU | |  | ATT no-waning | |  | Incremental | | |
| --- | --- | --- | --- | --- | --- | --- | --- | --- | --- |
|  | QALY | Cost |  | QALY | Cost |  | QALY | Cost | ICER |
| Treatment duration of 0.5 years | 5.3 | 353 |  | 6.0 | 365 |  | 0.75 | 11 | 15 |
| Continuous treatment until moderate dementia (MMSE <20) | 5.3 | 353 |  | 5.9 | 496 |  | 0.62 | 142 | 230 |
| Slowing of cognitive decline: 15% | 5.3 | 353 |  | 5.6 | 388 |  | 0.34 | 34 | 100 |
| Slowing of cognitive decline: 45% | 5.3 | 353 |  | 6.4 | 373 |  | 1.14 | 19 | 17 |
| Treatment effect stops at moderate dementia (MMSE <20) | 5.3 | 353 |  | 5.9 | 385 |  | 0.61 | 32 | 52 |
| Voluntary treatment discontinuation of 10%/yr | 5.3 | 353 |  | 5.9 | 382 |  | 0.63 | 29 | 46 |

*All scenarios were deviations from the of the 0% waning base case scenario. Costs are presented per 1000 euros.*

*Abbreviations: CAU = Care as usual, ATT = amyloid-targeting therapies, QALY = quality-adjusted life years, ICER = incremental cost-effectiveness ratio, MMSE = mini-mental state examination.*

**Supplemental Table 6:** ICERs for the scenarios after changing the treatment population, cost or discounting rate

| Scenario | CAU | |  | ATT no-waning | |  | Incremental | | |
| --- | --- | --- | --- | --- | --- | --- | --- | --- | --- |
|  | QALY | Cost |  | QALY | Cost |  | QALY | Cost | Icer |
| Simulation starts in mild dementia in the community (MMSE >20) | 3.3 | 393 |  | 3.8 | 425 |  | 0.51 | 31 | 62 |
| Simulation starts in MCI | 5.4 | 350 |  | 6.2 | 378 |  | 0.74 | 28 | 38 |
| No cost of screening for ATT eligibility | 5.3 | 353 |  | 6.0 | 379 |  | 0.72 | 25 | 36 |
| Not including infusion costs (analogous to using subcutaneous ATTs) | 5.3 | 353 |  | 6.0 | 379 |  | 0.72 | 25 | 36 |
| Yearly cost and utility discounting: 0% | 5.6 | 451 |  | 6.5 | 487 |  | 0.83 | 36 | 43 |
| Yearly cost and utility discounting: 5% | 4.5 | 303 |  | 5.1 | 329 |  | 0.51 | 26 | 51 |
| Healthcare perspective | 5.8 | 313 |  | 6.7 | 331 |  | 0.66 | 22 | 33 |

*All scenarios were deviations from the of the 0% waning base case scenario. Costs are presented per 1000 euros.*

*Abbreviations: CAU = Care as usual, ATT = amyloid-targeting therapies, QALY = quality-adjusted life years, ICER = incremental cost-effectiveness ratio, MMSE = mini-mental state examination.*

**Supplemental Table 7:** Cost and utilities for the three investigated treatment mechanisms

| Treatment effect | CAU | |  | ATT | |  | Incremental | | |
| --- | --- | --- | --- | --- | --- | --- | --- | --- | --- |
|  |  | |  | 0% waning/year | |  |  |  |  |
|  | QALY | Cost |  | QALY | Cost |  | QALY | Cost | ICER |
| 30% slowing cognitive decline (leaky) | 5.3 | 353 |  | 6.0 | 382 |  | 0.72 | 29 | 40 |
| 30% slowing cognitive decline (all-or-nothing) | 5.3 | 353 |  | 6.0 | 373 |  | 0.79 | 19 | 25 |
| 30% hazard reduction (leaky) | 5.2 | 356 |  | 6.3 | 400 |  | 1.08 | 44 | 41 |
|  |  |  |  | 20% waning/year | |  | | | |
| 30% slowing cognitive decline (leaky) | 5.3 | 353 |  | 5.5 | 391 |  | 0.25 | 37 | 151 |
| 30% slowing cognitive decline (all-or-nothing) | 5.3 | 353 |  | 5.5 | 392 |  | 0.24 | 38 | 162 |
| 30% hazard reduction (leaky) | 5.2 | 356 |  | 5.6 | 394 |  | 0.46 | 39 | 83 |

*Costs are presented per 1000 euros. Abbreviations: CAU = Care as usual, ATT = amyloid-targeting therapies, ICER = incremental cost-effectiveness ratio, MMSE = mini-mental state examination.*

**Supplemental Table 8**: Weibull hazard model for the transition from MCI to dementia

|  | Coefficient (standard error) |
| --- | --- |
| Shape | 0.29 (0.05) |
| Scale intercept | 0.99 (0.49) |
| Age at baseline, yrs | 0.005 (0.007) |
| Sex, female as reference | 0.17 (0.10) |

*Weibull parametrisation is the same as the rweibull function in R and are presented on a log-scale.^14^ Reducing the scale shifts the distribution towards earlier occurrence of outcomes*

**Supplemental Table 9**: Weibull hazard model for the transition from dementia to institution or death outside of community

|  | Coefficient (standard error) |
| --- | --- |
| Shape | 0.52 (0.04) |
| Scale intercept | 2.80 (0.31) |
| Institutionalisation as outcome, death in community as reference | -0.64 (0.07) |
| Sex, female reference | -0.20 (0.06) |
| Age at dementia diagnosis, yrs | -0.003 (0.004) |
| Lived with care partner at baseline, living alone reference | 0.10 (0.07) |
| Diagnosis of dementia at baseline, MCI as reference | -0.16 (0.11) |

*Weibull parametrisation is the same as the rweibull function in R and are presented on a log-scale.^14^ Reducing the scale shifts the distribution towards earlier occurrence of outcomes*

**Supplemental Table** **10**: Weibull model for transition from institutionalisation to death

|  | Coefficient (standard error) |
| --- | --- |
| Shape | 0.07 (0.07) |
| Scale intercept | 3.46 (0.73) |
| Sex, female reference | -0.23 (0.16) |
| Age at institutionalisation, yrs | -0.02 (0.01) |
| Lived with care partner at baseline, living alone reference | -0.87 (0.23) |

*Weibull parametrisation is the same as the rweibull function in R and are presented on a log-scale.^14^ Reducing the scale shifts the distribution towards earlier occurrence of outcomes. This table was copied from Veere et al. (2026).^15^*

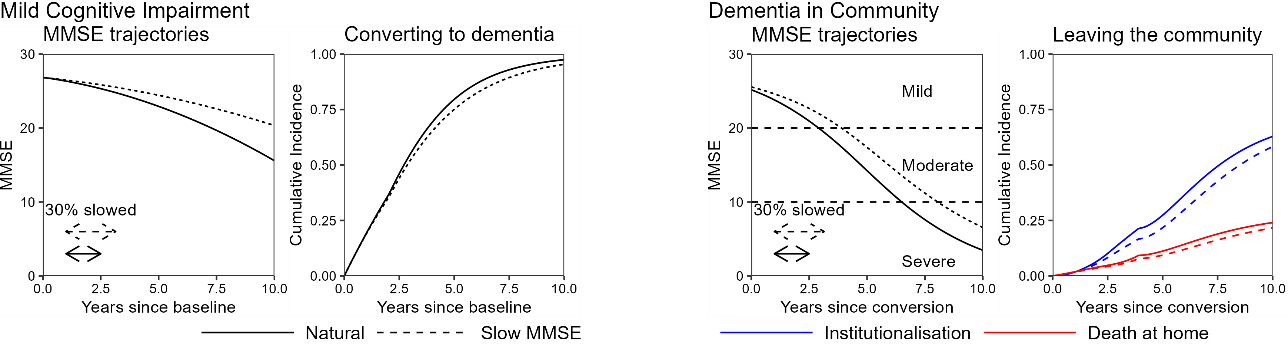

**Supplemental Figure 1**: Illustration of the treatment effect

*This figure illustrates a 30% slowing of MMSE in the MCI and mild dementia states, and its subsequent effect on the cumulative incidence functions. This figured was copied from Veere et al. (2026).^15^*

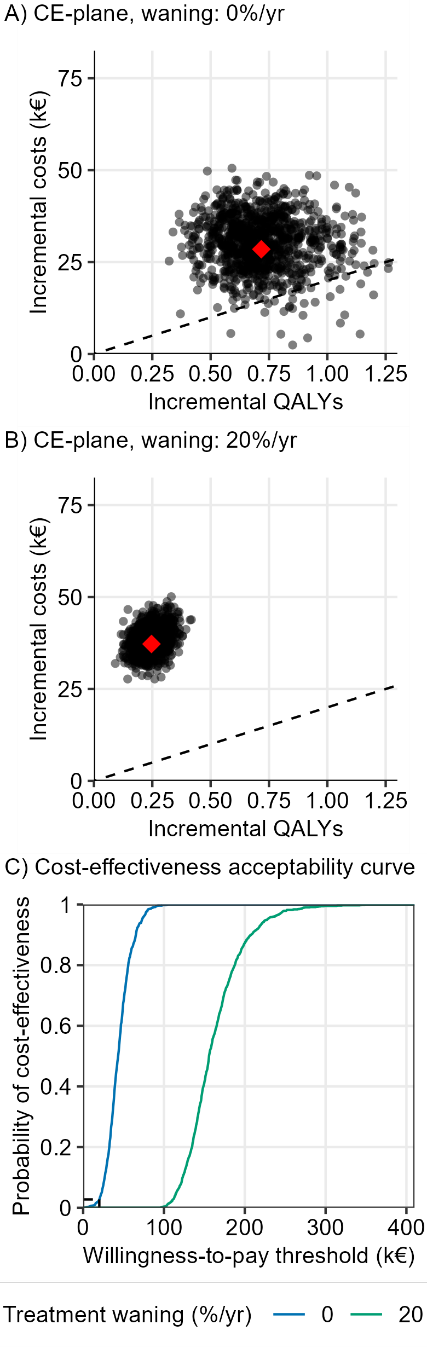

**Supplemental Figure 2**: Cost-effectiveness planes and cost-effectiveness acceptability curve of the base cases

Each dot in the cost-effectiveness plane symbolises the results of one set of model parameters. The dotted lines represent a €20,000/QALY willingness-to-pay threshold.

Abbreviations: CE= cost-effectiveness plane. QALY = quality adjusted life year.
